# A Large-Scale Clinical Validation Study Using nCapp Cloud Plus Terminal by Frontline Doctors for the Rapid Diagnosis of COVID-19 and COVID-19 pneumonia in China

**DOI:** 10.1101/2020.08.07.20163402

**Authors:** Dawei Yang, Tao Xu, Xun Wang, Deng Chen, Ziqiang Zhang, Lichuan Zhang, Jie Liu, Kui Xiao, Li Bai, Yong Zhang, Lin Zhao, Lin Tong, Chaomin Wu, Yaoli Wang, Chunling Dong, Maosong Ye, Yu Xu, Zhenju Song, Hong Chen, Jing Li, Jiwei Wang, Fei Tan, Hai Yu, Jian Zhou, Jinming Yu, Chunhua Du, Hongqing Zhao, Yu Shang, Linian Huang, Jianping Zhao, Yang Jin, Charles A. Powell, Yuanlin Song, Chunxue Bai

## Abstract

**Background:** The outbreak of coronavirus disease 2019 (COVID-19) has become a global pandemic acute infectious disease, especially with the features of possible asymptomatic carriers and high contagiousness. It causes acute respiratory distress syndrome and results in a high mortality rate if pneumonia is involved. Currently, it is difficult to quickly identify asymptomatic cases or COVID-19 patients with pneumonia due to limited access to reverse transcription-polymerase chain reaction (RT-PCR) nucleic acid tests and CT scans, which facilitates the spread of the disease at the community level, and contributes to the overwhelming of medical resources in intensive care units.

**Goal:** This study aimed to develop a scientific and rigorous clinical diagnostic tool for the rapid prediction of COVID-19 cases based on a COVID-19 clinical case database in China, and to assist global frontline doctors to efficiently and precisely diagnose asymptomatic COVID-19 patients and cases who had a false-negative RT-PCR test result.

**Methods:** With online consent, and the approval of the ethics committee of Zhongshan Hospital Fudan Unversity (approval number B2020-032R) to ensure that patient privacy is protected, clinical information has been uploaded in real-time through the New Coronavirus Intelligent Auto-diagnostic Assistant Application of cloud plus terminal (nCapp) by doctors from different cities (Wuhan, Shanghai, Harbin, Dalian, Wuxi, Qingdao, Rizhao, and Bengbu) during the COVID-19 outbreak in China. By quality control and data anonymization on the platform, a total of 3,249 cases from COVID-19 high-risk groups were collected. These patients had SARS-CoV-2 RT-PCR test results and chest CT scans, both of which were used as the gold standard for the diagnosis of COVID-19 and COVID-19 pneumonia. In particular, the dataset included 137 indeterminate cases who initially did not have RT-PCR tests and subsequently had positive RT-PCR results, 62 suspected cases who initially had false-negative RT-PCR test results and subsequently had positive RT-PCR results, and 122 asymptomatic cases who had positive RT-PCR test results, amongst whom 31 cases were diagnosed. We also integrated the function of a survey in nCapp to collect user feedback from frontline doctors.

**Findings:** We applied the statistical method of a multi-factor regression model to the training dataset (1,624 cases) and developed a prediction model for COVID-19 with 9 clinical indicators that are fast and accessible: ‘Residing or visiting history in epidemic regions’, ‘Exposure history to COVID-19 patient’, ‘Dry cough’, ‘Fatigue’, ‘Breathlessness’, ‘No body temperature decrease after antibiotic treatment’, ‘Fingertip blood oxygen saturation ≤93%’, ‘Lymphopenia’, and ‘C-reactive protein (CRP) increased’. The area under the receiver operating characteristic (ROC) curve (AUC) for the model was 0.88 (95% CI: 0.86, 0.89) in the training dataset and 0.84 (95% CI: 0.82, 0.86) in the validation dataset (1,625 cases). To ensure the sensitivity of the model, we used a cutoff value of 0.09. The sensitivity and specificity of the model were 98.0% (95% CI: 96.9%, 99.1%) and 17.3% (95% CI: 15.0%, 19.6%), respectively, in the training dataset, and 96.5% (95% CI: 95.1%, 98.0%) and 18.8% (95% CI: 16.4%, 21.2%), respectively, in the validation dataset. In the subset of the 137 indeterminate cases who initially did not have RT-PCR tests and subsequently had positive RT-PCR results, the model predicted 132 cases, accounting for 96.4% (95% CI: 91.7%, 98.8%) of the cases. In the subset of the 62 suspected cases who initially had false-negative RT-PCR test results and subsequently had positive RT-PCR results, the model predicted 59 cases, accounting for 95.2% (95% CI: 86.5%, 99.0%) of the cases. Considering the specificity of the model, we used a cutoff value of 0.32. The sensitivity and specificity of the model were 83.5% (95% CI: 80.5%, 86.4%) and 83.2% (95% CI: 80.9%, 85.5%), respectively, in the training dataset, and 79.6% (95% CI: 76.4%, 82.8%) and 81.3% (95% CI: 78.9%, 83.7%), respectively, in the validation dataset, which is very close to the published AI model.

The results of the online survey ‘Questionnaire Star’ showed that 90.9% of nCapp users in WeChat mini programs were ‘satisfied’ or ‘very satisfied’ with the tool. The WeChat mini program received a significantly higher satisfaction rate than other platforms, especially for ‘availability and sharing convenience of the App’ and ‘fast speed of log-in and data entry’.

**Discussion:** With the assistance of nCapp, a mobile-based diagnostic tool developed from a large database that we collected from COVID-19 high-risk groups in China, frontline doctors can rapidly identify asymptomatic patients and avoid misdiagnoses of cases with false-negative RT-PCR results. These patients require timely isolation or close medical supervision. By applying the model, medical resources can be allocated more reasonably, and missed diagnoses can be reduced. In addition, further education and interaction among medical professionals can improve the diagnostic efficiency for COVID-19, thus avoiding the transmission of the disease from asymptomatic patients at the community level.

## Introduction

The first outbreak of coronavirus disease 2019 (COVID-19) was recorded at the end of 2019 in Wuhan city, Hubei Province, China, and currently more than 185 countries and territories have reported COVID-19 cases.^1^ According to the latest statistics, more than 4.02 million cases have been diagnosed worldwide, with the majority of cases now in the U.S.^2^ To prevent and control COVID-19, and to halt the spread of the disease as quickly as possible, it is crucial to provide early detection, timely diagnosis, and proper management of COVID-19 cases. However, the clinical manifestations of COVID-19 vary at different disease stages, and the sensitivity of viral nucleic acid diagnostic tests cannot ensure 100% identification of infected patients. Therefore, chest radiology was added to the diagnostic standard in the fifth edition of the *Chinese COVID-19 diagnosis and treatment guideline* (‘Chinese Guideline’).^3^ In the space of just two months, the Chinese Guideline has been updated to the seventh edition^4^ to provide better guidance for doctors in the diagnosis and treatment of COVID-19.

The wide spread of COVID-19 and large number of newly infected patients require doctors from different specialties (including general practitioners [GPs]) to participate in the diagnosis, treatment, and management of the disease. The earliest guidelines from the World Health Organization (WHO)^5^ and China^4^ listed multiple clinical manifestations and laboratory tests. In combination with expert opinion, there were 20 diagnosis-related factors in total, which made it difficult for non-expert doctors to decide which ones were the most important determinants. As a result, there was a long learning curve for frontline doctors to acquire the skills for the precise and timely diagnosis of the disease. The complexity of diagnosis led to missed diagnoses or delayed diagnoses, and to some extent, facilitated the spread of COVID-19 at the community level.^6^ Additionally, due to the lack of a modern and standardized medical quality control system, there were missed diagnoses or misdiagnoses, especially among patients with false-negative nucleic acid test results or false-negative CT scan results.^7^ Currently, there is no rapid clinical diagnosis model specially designed for COVID-19 based on clinical informatics. To create such a model, it is necessary to optimize and screen the essential diagnostic determinants from the existing diagnosis-related factors, and to assist clinical practice by applying user-friendly internet tools.

To address this gap, we developed the New Coronavirus Intelligent Auto-diagnostic Assistant Application of cloud plus terminal (nCapp), a mobile-based tool as a mini program within WeChat (a social media App), exploiting the fact that almost all doctors in China are smartphone users. We conducted a prospective clinical study (ClinicalTrials.gov registration number NCT04275947). We aimed to apply nCapp, with the assistance of modern statistical methods, to optimize and select a group of essential diagnostic determinants for COVID-19, with high sensitivity and specificity, and to support doctors in their clinical practice, facilitating better management of COVID-19 patients.

## Methods

### Basics of nCapp

This study applied nCapp, a mobile-based diagnostic tool, to conduct a multi-center clinical study.^8^ In addition to following the standards for designing a mobile App, the back-end was developed using Java, and Object Storage Service (OSS) was used for data storage (Figure 1). nCapp applies statistical methods to optimize and select the essential diagnostic determinants for COVID-19. It also provides an easy and accessible intelligent process system to assist doctors from all specialties in their clinical practice. To better describe its essence and function, we use the name ‘nCapp Cloud Plus Terminal’.^9^ It is the first global mobile application developed for the early diagnosis of infectious diseases, especially COVID-19. To be specific, cases are classified into different risk stratification categories, in line with the *Expert consensus on COVID-19 pneumonia diagnosis and treatment using Intelligent Auto-Diagnostic Assistant Application (nCappf* and differentiated interventions are tiered to each category. For example, interventions for critical cases include a cloud link to senior doctors, symptomatic treatment, respiratory support, circulation support, psychological guidance, and traditional Chinese medicine (TCM) treatment; interventions for suspected cases include reporting or transfer to a superior hospital, a cloud link to senior doctors, and continuation of the current treatment; interventions for indeterminate cases include a cloud link to senior doctors, continuation of the current treatment, and isolation and observation for 14 days.

**Figure 1:**
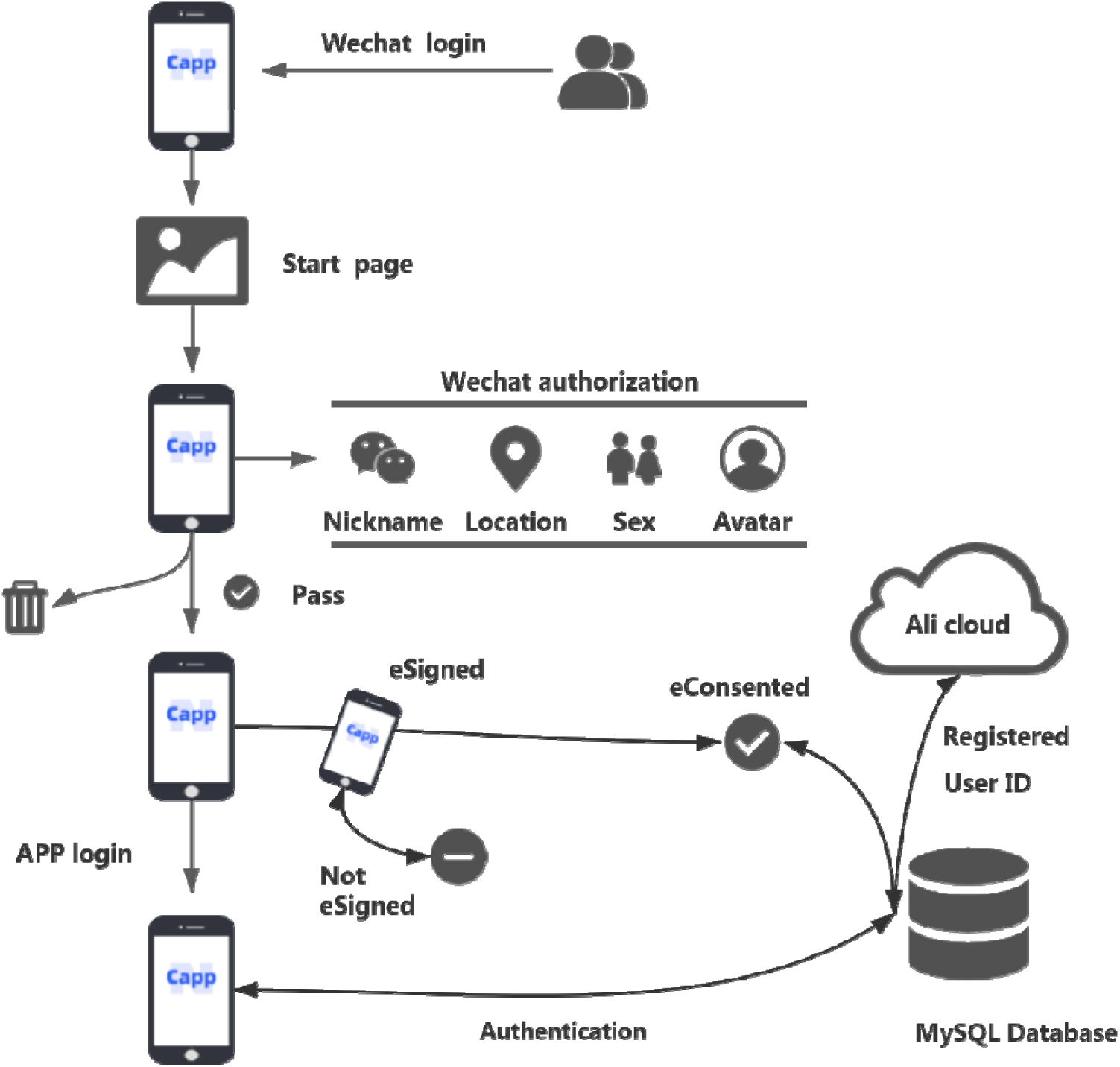
Flow chart of data information processing through the nCapp Cloud Plus Terminal model

### Data collection

The nCapp Cloud Plus Terminal model can be used for many diseases and public health events. It has both Chinese and English versions, and it can be accessed by both iPhone and Android users. The users, who are frontline doctors participating in this study, first need to search for ‘WeChat’ and install the WeChat App on their phones (iPhone users install through the ‘AppStore’; Android phone users install through the ‘PlayStore’). The users then follow the instructions in WeChat to register an account and use the ‘scan’ function to scan the QR code in Figure S1 to start nCapp. Patient privacy is protected with the required online consent and the study protocol was approved by the ethics committee of Zhongshan Hospital Fudan University (approval number B2020-032R). Since the start of the study on February 13, 2020, clinical information on 3,249 cases from COVID-19 high-risk groups has been uploaded through nCapp, with their SARS-CoV-2 reverse transcription-polymerase chain reaction (RT-PCR) nucleic acid test results and chest CT scan results available. The training dataset of the statistical model randomly included 1,624 cases, and the remaining 1,625 cases were included in the validation dataset. The patients were of COVID-19 high-risk groups from Wuhan, Shanghai, Harbin, Dalian, Wuxi, Qingdao, Rizhao, and Bengbu, who were admitted to local infectious disease hospitals. All data were recorded anonymously and uploaded by local doctors.

### Statistical methods

The data on 3,249 COVID-19 patients uploaded through nCapp were input into the dataset. We applied stratified random sampling to sample 1,624 patients for the training dataset of the statistical model, and the remaining 1,625 cases were included in the validation dataset (i.e. all the samples were first divided into two strata based on the RT-PCR results; 606 cases with positive RT-PCR results and 1,018 cases with negative RT-PCR results were randomly included in the training dataset, and the remaining 1,625 cases were included in the validation dataset). SAS 9.4 and R 3.6.2 statistical software packages were used for data analysis and plotting. Odds ratios (ORs) and 95% confidence intervals (CIs) (as the approximate estimate of the relative risk) were reported to estimate the strength of association between each variable and the test result. Single-factor logistic regression analysis was first applied to the training dataset. The RT-PCR test result was entered into the model as a dependent variable (Y: negative=0, positive=1). The effects of different diagnostic factors were ranked based on the results from a single factor analysis, with 0.05 as the significance level for factor inclusion and 0.1 as the significance level for factor exclusion. Independent variables were selected using the step-forward method of partial maximum likelihood estimation. Then stepwise multi-factor logistic regression analysis was applied to obtain the probability model for cases with positive test results (i.e. COVID-19 patients). The goodness of fit of the model on the training dataset and the validation dataset was used to measure the prediction accuracy.

## Results

### Demographics

Table 1 demonstrates that the basic demographic characteristics of the cases included in this study approximately coincide with the clinical characteristics of COVID-19 patients in China (Guan et al.), which certifies the reliability of the samples and the generalizability of this research.

**Table 1:**
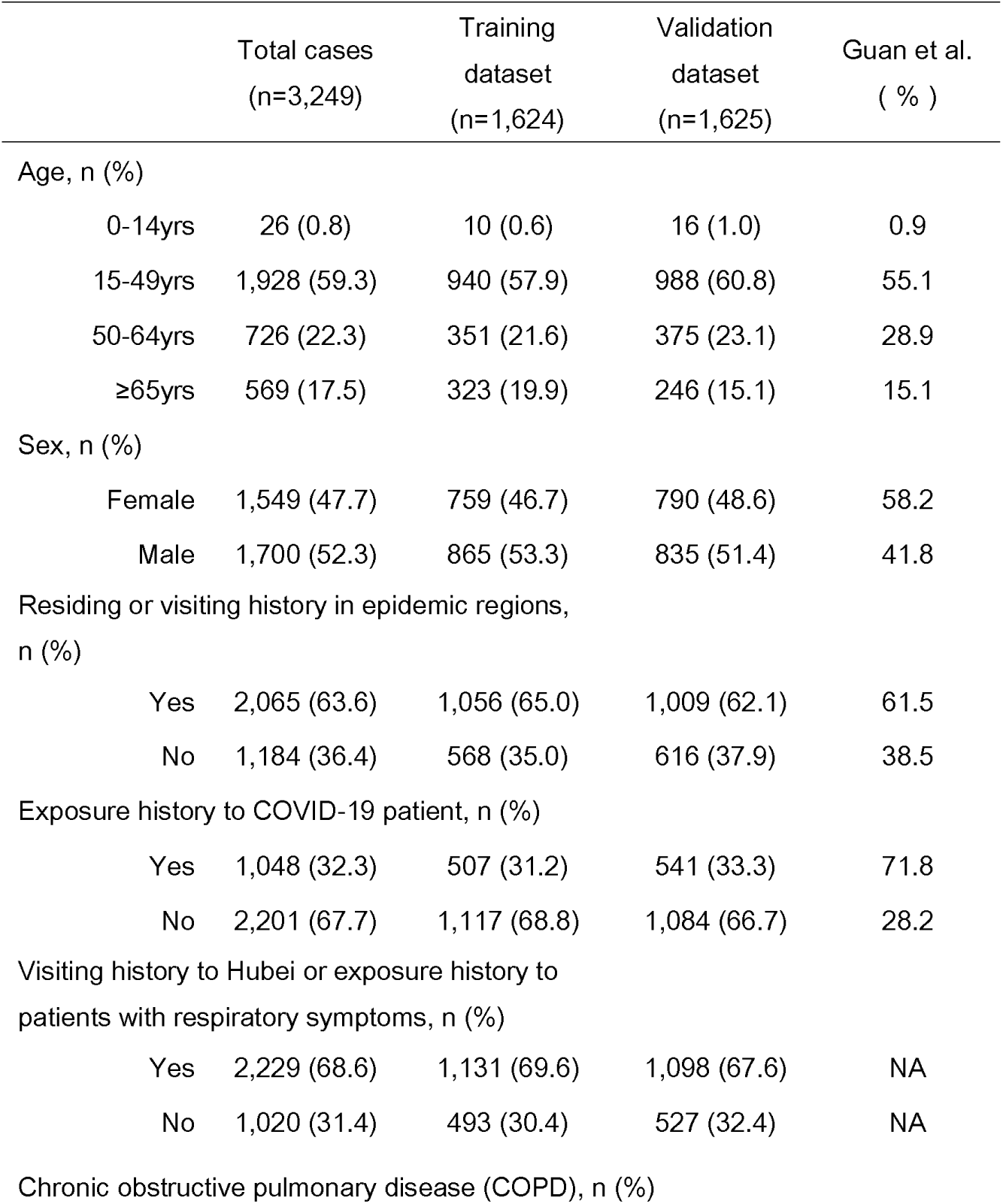
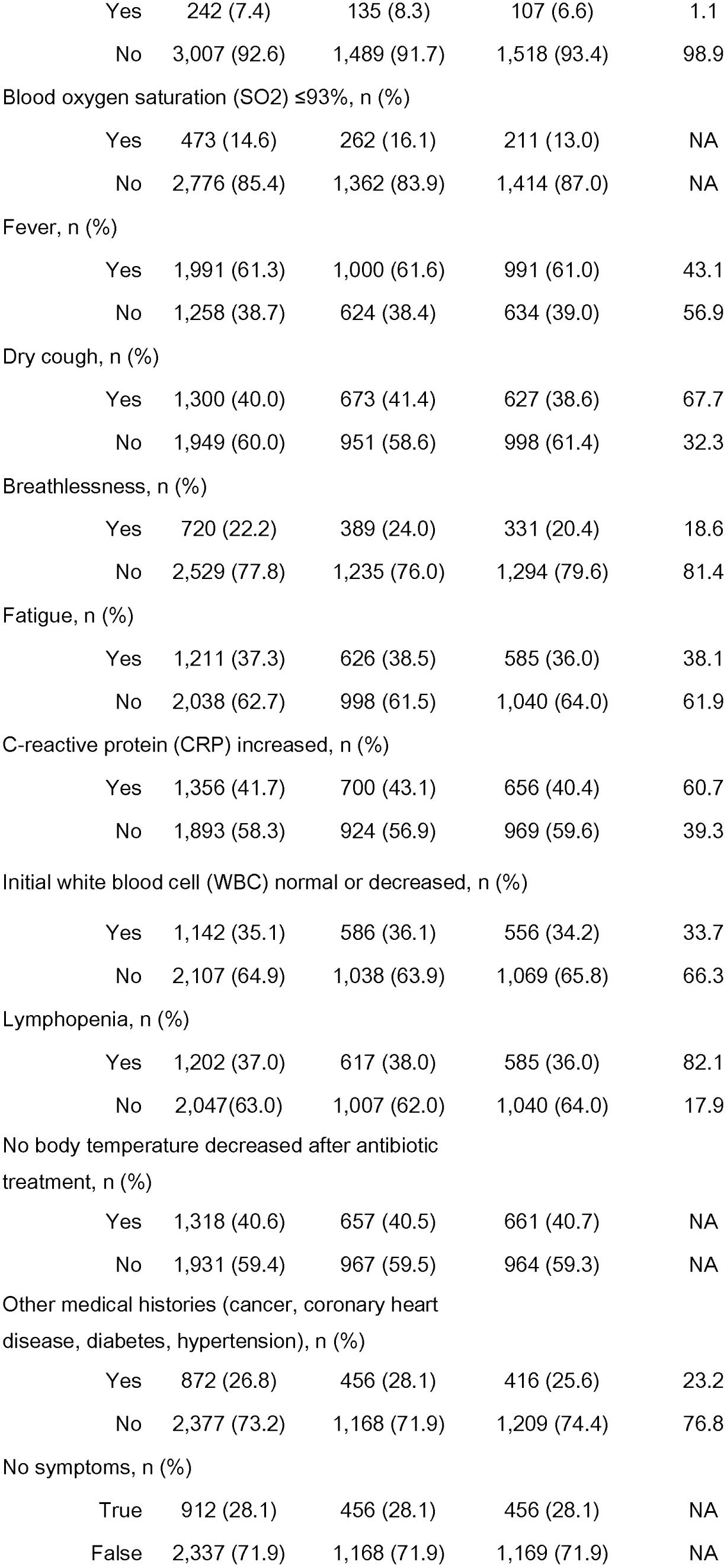
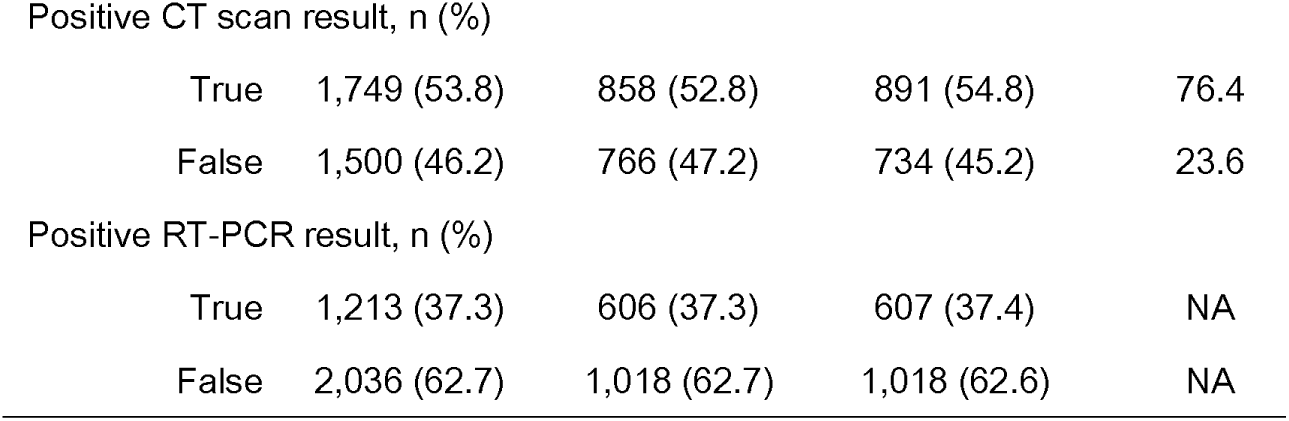
Demographics of the training dataset and the validation dataset

|  | Total cases<br>(n=3,249) | Training<br>dataset<br>(n=1,624) | Validation<br>dataset<br>(n=1,625) | Guan et al.<br>( % ) |
| --- | --- | --- | --- | --- |
| Age, n (%) |  |  |  |  |
| 0-14yrs | 26 (0.8) | 10 (0.6) | 16 (1.0) | 0.9 |
| 15-49yrs | 1,928 (59.3) | 940 (57.9) | 988 (60.8) | 55.1 |
| 50-64yrs | 726 (22.3) | 351 (21.6) | 375 (23.1) | 28.9 |
| ≥65yrs | 569 (17.5) | 323 (19.9) | 246 (15.1) | 15.1 |
| Sex, n (%) |  |  |  |  |
| Female | 1,549 (47.7) | 759 (46.7) | 790 (48.6) | 58.2 |
| Male | 1,700 (52.3) | 865 (53.3) | 835 (51.4) | 41.8 |
| Residing or visiting history in epidemic regions,<br>n (%) |  |  |  |  |
| Yes | 2,065 (63.6) | 1,056 (65.0) | 1,009 (62.1) | 61.5 |
| No | 1,184 (36.4) | 568 (35.0) | 616 (37.9) | 38.5 |
| Exposure history to COVID-19 patient, n (%) |  |  |  |  |
| Yes | 1,048 (32.3) | 507 (31.2) | 541 (33.3) | 71.8 |
| No | 2,201 (67.7) | 1,117 (68.8) | 1,084 (66.7) | 28.2 |
| Visiting history to Hubei or exposure history to<br>patients with respiratory symptoms, n (%) |  |  |  |  |
| Yes | 2,229 (68.6) | 1,131 (69.6) | 1,098 (67.6) | NA |
| No | 1,020 (31.4) | 493 (30.4) | 527 (32.4) | NA |
| Chronic obstructive pulmonary disease (COPD), n (%) |  |  |  |  |
| Yes | 242 (7.4) | 135 (8.3) | 107 (6.6) | 1.1 |
| No | 3,007 (92.6) | 1,489 (91.7) | 1,518 (93.4) | 98.9 |
| Blood oxygen saturation (SO<sub>2</sub>) ≤93%, n (%) |  |  |  |  |
| Yes | 473 (14.6) | 262 (16.1) | 211 (13.0) | NA |
| No | 2,776 (85.4) | 1,362 (83.9) | 1,414 (87.0) | NA |
| Fever, n (%) |  |  |  |  |
| Yes | 1,991 (61.3) | 1,000 (61.6) | 991 (61.0) | 43.1 |
| No | 1,258 (38.7) | 624 (38.4) | 634 (39.0) | 56.9 |
| Dry cough, n (%) |  |  |  |  |
| Yes | 1,300 (40.0) | 673 (41.4) | 627 (38.6) | 67.7 |
| No | 1,949 (60.0) | 951 (58.6) | 998 (61.4) | 32.3 |
| Breathlessness, n (%) |  |  |  |  |
| Yes | 720 (22.2) | 389 (24.0) | 331 (20.4) | 18.6 |
| No | 2,529 (77.8) | 1,235 (76.0) | 1,294 (79.6) | 81.4 |
| Fatigue, n (%) |  |  |  |  |
| Yes | 1,211 (37.3) | 626 (38.5) | 585 (36.0) | 38.1 |
| No | 2,038 (62.7) | 998 (61.5) | 1,040 (64.0) | 61.9 |
| C-reactive protein (CRP) increased, n (%) |  |  |  |  |
| Yes | 1,356 (41.7) | 700 (43.1) | 656 (40.4) | 60.7 |
| No | 1,893 (58.3) | 924 (56.9) | 969 (59.6) | 39.3 |
| Initial white blood cell (WBC) normal or decreased, n (%) |  |  |  |  |
| Yes | 1,142 (35.1) | 586 (36.1) | 556 (34.2) | 33.7 |
| No | 2,107 (64.9) | 1,038 (63.9) | 1,069 (65.8) | 66.3 |
| Lymphopenia, n (%) |  |  |  |  |
| Yes | 1,202 (37.0) | 617 (38.0) | 585 (36.0) | 82.1 |
| No | 2,047 (63.0) | 1,007 (62.0) | 1,040 (64.0) | 17.9 |
| No body temperature decreased after antibiotic treatment, n (%) |  |  |  |  |
| Yes | 1,318 (40.6) | 657 (40.5) | 661 (40.7) | NA |
| No | 1,931 (59.4) | 967 (59.5) | 964 (59.3) | NA |
| Other medical histories (cancer, coronary heart disease, diabetes, hypertension), n (%) |  |  |  |  |
| Yes | 872 (26.8) | 456 (28.1) | 416 (25.6) | 23.2 |
| No | 2,377 (73.2) | 1,168 (71.9) | 1,209 (74.4) | 76.8 |
| No symptoms, n (%) |  |  |  |  |
| True | 912 (28.1) | 456 (28.1) | 456 (28.1) | NA |
| False | 2,337 (71.9) | 1,168 (71.9) | 1,169 (71.9) | NA |
| Positive CT scan result, n (%) |  |  |  |  |
| True | 1,749 (53.8) | 858 (52.8) | 891 (54.8) | 76.4 |
| False | 1,500 (46.2) | 766 (47.2) | 734 (45.2) | 23.6 |
| Positive RT-PCR result, n (%) |  |  |  |  |
| True | 1,213 (37.3) | 606 (37.3) | 607 (37.4) | NA |
| False | 2,036 (62.7) | 1,018 (62.7) | 1,018 (62.6) | NA |

Table 2 presents the comparison of demographic indicators among four subsets: cases with positive RT-PCR and CT scan results, cases with a positive RT-PCR result and negative CT scan result, cases with a negative RT-PCR result and positive CT scan result, and cases with negative RT-PCR and CT scan results. It shows statistically significant differences in all the demographic indicators (*P*<0.001), except for sex.

**Table 2:**
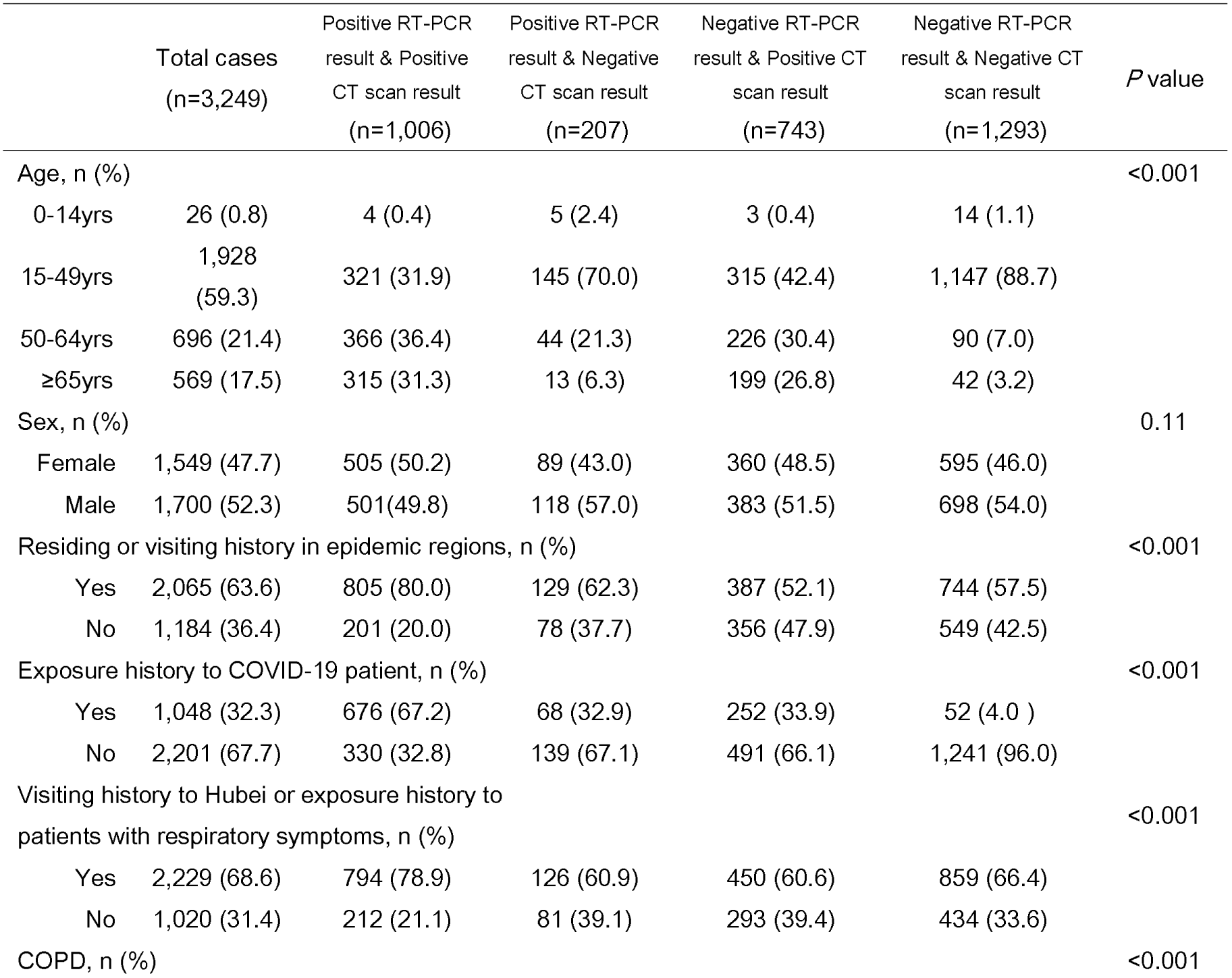
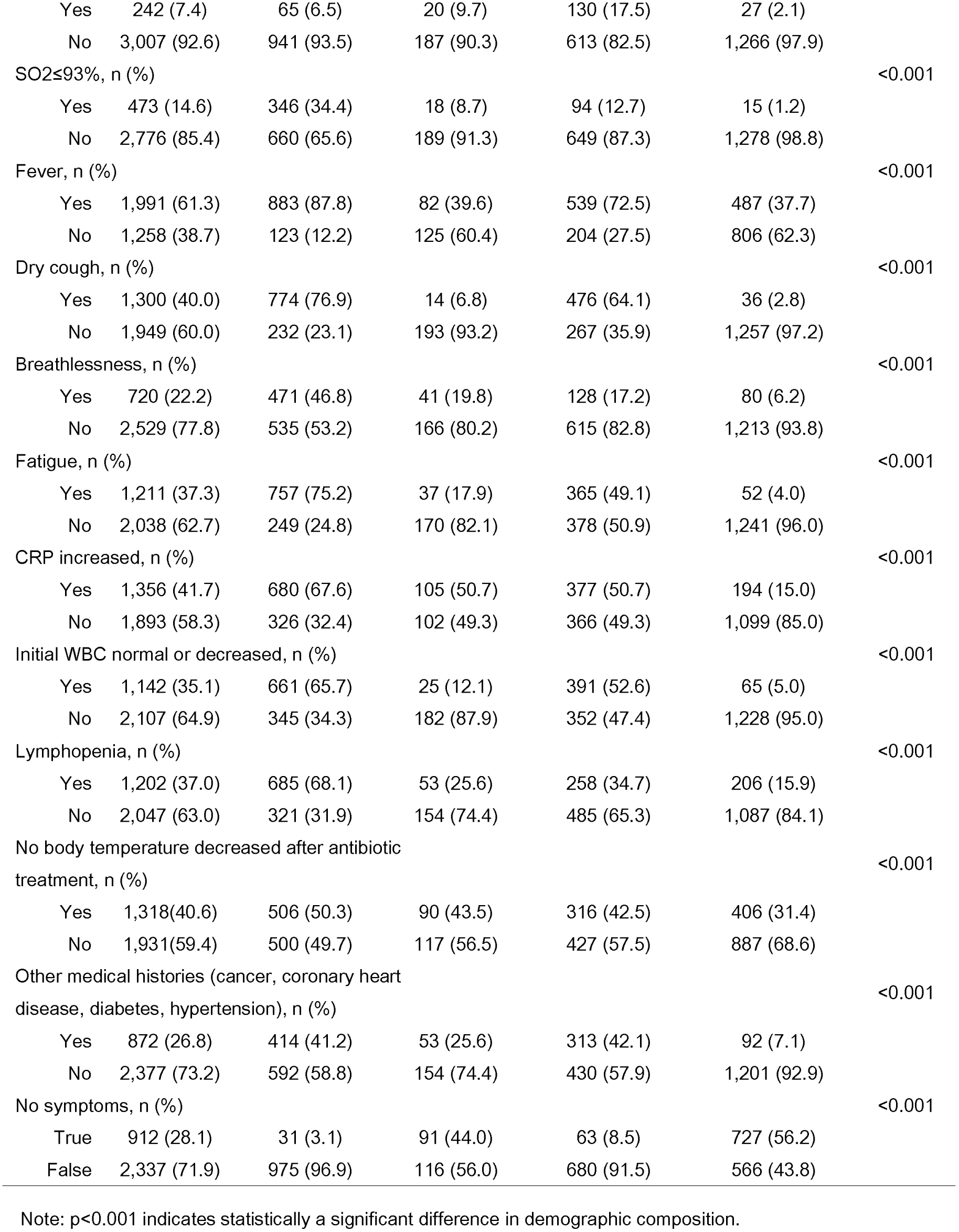
Demographics of the four subsets

### Case composition and factors affecting the four subsets

#### Composition of all the cases

As illustrated in Figure 2, among the asymptomatic patients (no dry cough, fever, breathlessness, or fatigue), cases with negative RT-PCR and CT scan results account for the vast majority (79.7%), and the other three subsets account for a low proportion. Among the symptomatic patients (at least one symptom among dry cough, fever, breathlessness, and fatigue), cases with positive RT-PCR and CT scan results account for the highest proportion (41.7%), followed by cases with negative RT-PCR result and positive CT scan result, and cases with negative RT-PCR and CT scan results (both over 20%), while cases with positive RT-PCR result and negative CT scan result account for the lowest proportion (5%). Combined with Figure 3, it shows that ‘no symptoms’ and ‘no exposure history to COVID-19 patient’ are sensitive assessment indicators for cases with negative RT-PCR and CT scan results; ‘with symptoms’ and ‘residing or visiting history in epidemic regions’ or ‘visiting history to Hubei or exposure history to patients with respiratory symptoms’ indicate a higher risk of cases with positive RT-PCR and CT scan results. It is also noteworthy that 13.4% of the asymptomatic cases have positive RT-PCR test results while 53.3% of the symptomatic cases have negative RT-PCR test results.

**Figure 2:**
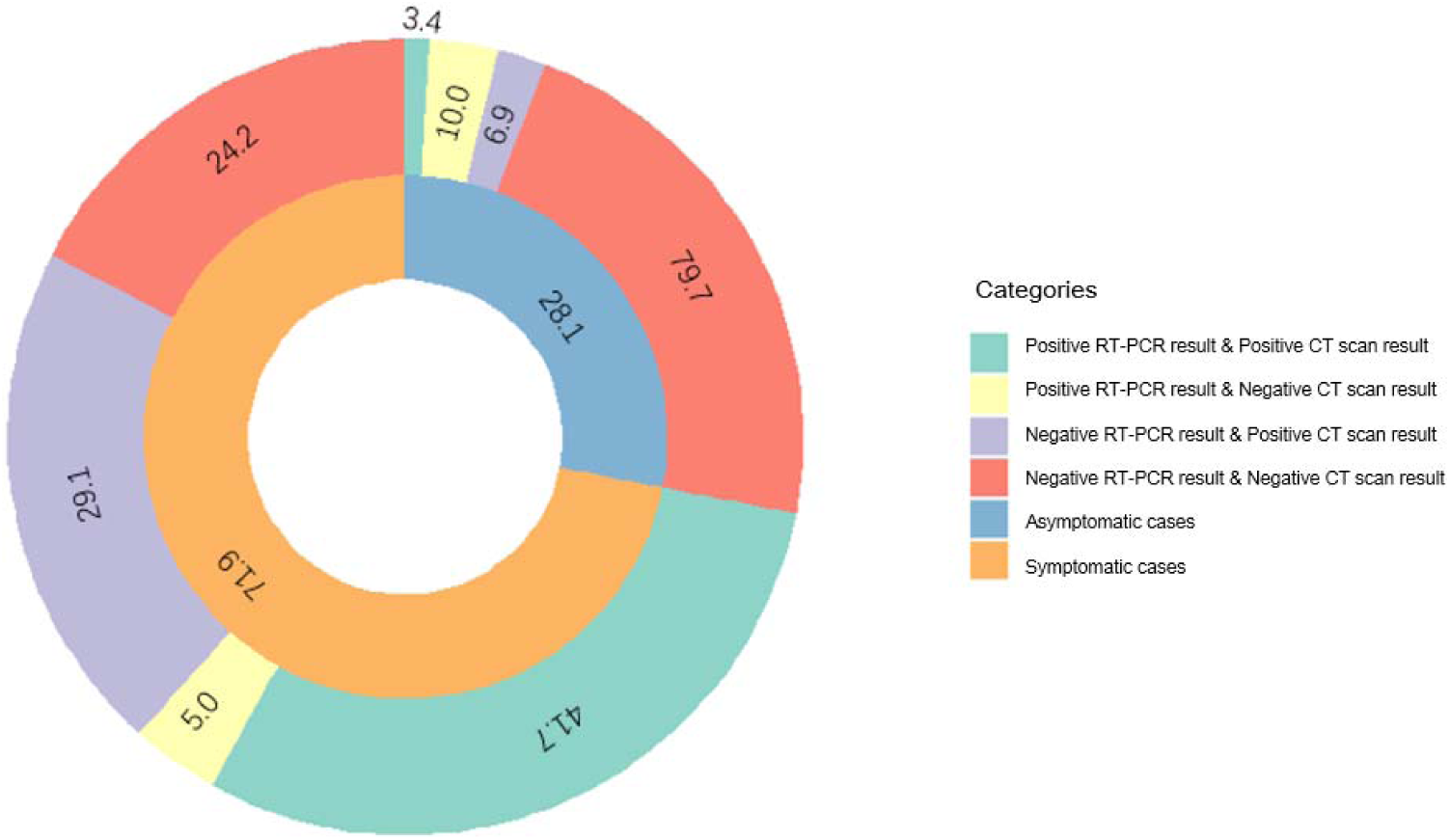
Composition of the cases (%)

**Figure 3:**
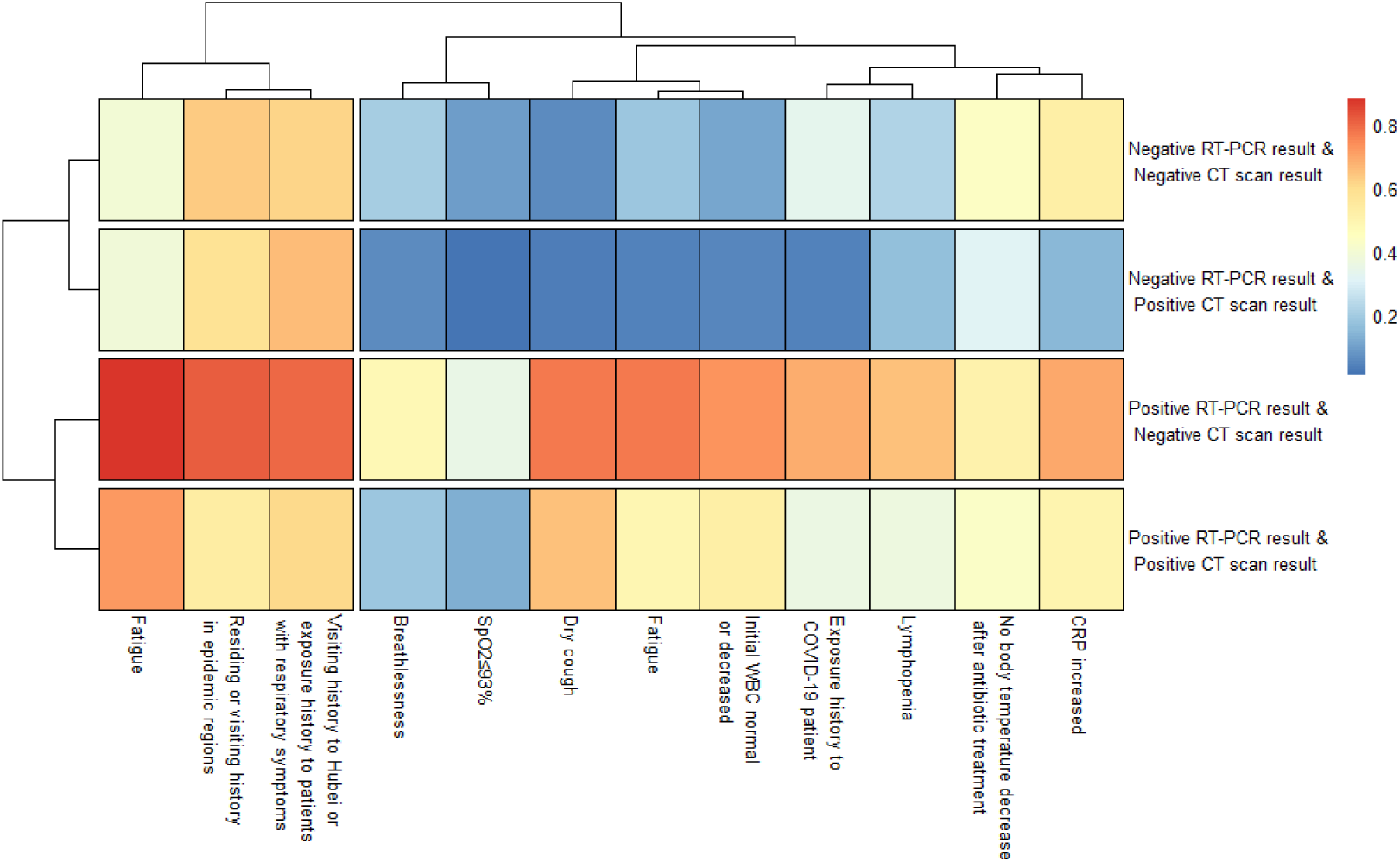
Clustering of the factors affecting the four subsets

### Probability model for COVID-19

Independent variables in the model of diagnostic factors were selected using the step-forward method of partial maximum likelihood estimation, with 0.05 as the significance level for factor inclusion, and 0.1 as the significance level for factor exclusion. After the single factor logistic regression analysis, a multi-factor regression analysis was applied, including ‘Residing or visiting history in epidemic regions’, ‘Exposure history to COVID-19 patient’, ‘Visiting history to Hubei or exposure history to patient with respiratory symptoms’, ‘Dry cough’, ‘Fever’, ‘No body temperature decrease after antibiotic treatment’, ‘Fatigue’, ‘Breathlessness’, ‘Fingertip blood oxygen saturation ≤93%’, ‘Lymphopenia’, and ‘CRP increased’. We obtained a probability model for cases with positive test results (i.e. COVID-19 patients). The equation expression for the model is:

Probability of COVID-19 = e^x^ / (1 + e^x^)

x = −3.13 + (0.99 * ‘Residing or visiting history in epidemic regions’) + (1.15 * ‘Exposure history to COVID-19 patient’) + (0.37 * ‘Dry cough’) + (0.52 * ‘Fatigue’) + (0.74 * ‘Breathlessness’) + (0.53 * ‘Fingertip blood oxygen saturation ≤93%’) + (0.58 * ‘Lymphopenia’) + (0.36 * ‘No body temperature decreased after antibiotic treatment’) + (1.10 * ‘CRP increased’). (Table 3)

**Table 3:** Probability model for COVID-19 based on logistic regression analysis

| | $\beta$ coefficient | SE | P-value | OR (95% CI) |
| --- | --- | --- | --- | --- |
| Intercept | -3.13 | 0.17 | <0.001 |  |
| Residing or visiting history in epidemic regions | 0.99 | 0.15 | <0.001 | 2.69(2.00,3.66) |
| Exposure history to COVID-19 patient | 1.15 | 0.16 | <0.001 | 3.16(2.29,4.34) |
| Dry cough | 0.37 | 0.17 | 0.03 | 1.45(1.04,2.02) |
| Fatigue | 0.52 | 0.17 | <0.001 | 1.68(1.20,2.36) |
| Breathlessness | 0.74 | 0.18 | <0.001 | 2.10(1.46,2.97) |
| SO<sub>2</sub>≤93% | 0.53 | 0.23 | 0.02 | 1.70(1.08,2.71) |
| Lymphopenia | 0.58 | 0.15 | <0.001 | 1.79(1.34,2.40) |
| No body temperature<br>decrease after antibiotic<br>treatment | 0.36 | 0.14 | 0.009 | 1.43(1.09,1.88) |
| CRP increased | 1.10 | 0.15 | <0.001 | 3.00(2.25,4.02) |

The AUC for the model was 0.88 (95% CI: 0.86, 0.89) in the training dataset (1,624 cases) and 0.84 (95% CI: 0.82, 0.86) in the validation dataset (1,625 cases). To ensure the sensitivity of the model, we used a cutoff value at 0.09. The sensitivity and specificity of the model were 98.0% (95% CI: 96.9%, 99.1%) and 17.3% (95% CI: 15.0%, 19.6%), respectively, in the training dataset, and 96.5% (95% CI: 95.1%, 98.0%) and 18.8% (95% CI: 16.4%, 21.2%), respectively, in the validation dataset. The model predicted 132 cases of the 137 indeterminate cases who initially did not have RT-PCR tests and subsequently had positive RT-PCR results, i.e. 96.4% (95% CI: 91.7%, 98.8%) in this subset, and 59 cases of the 62 suspected cases who initially had false-negative RT-PCR test results and subsequently had positive RT-PCR results, i.e. 95.2% (95% CI: 86.5%, 99.0%) in this subset. Considering the specificity of the model, we used a cutoff value at 0.32. The sensitivity and specificity of the model were 83.5% (95% CI: 80.5%, 86.4%) and 83.2% (95% CI: 80.9%, 85.5%), respectively, in the training dataset, and 79.6% (95% CI: 76.4%, 82.8%) and 81.3% (95% CI: 78.9%, 83.7%), respectively, in the validation dataset, which is very close to the published AI model (sensitivity:84.3%, specificity:82.8%).^10^

**Figure 4:**
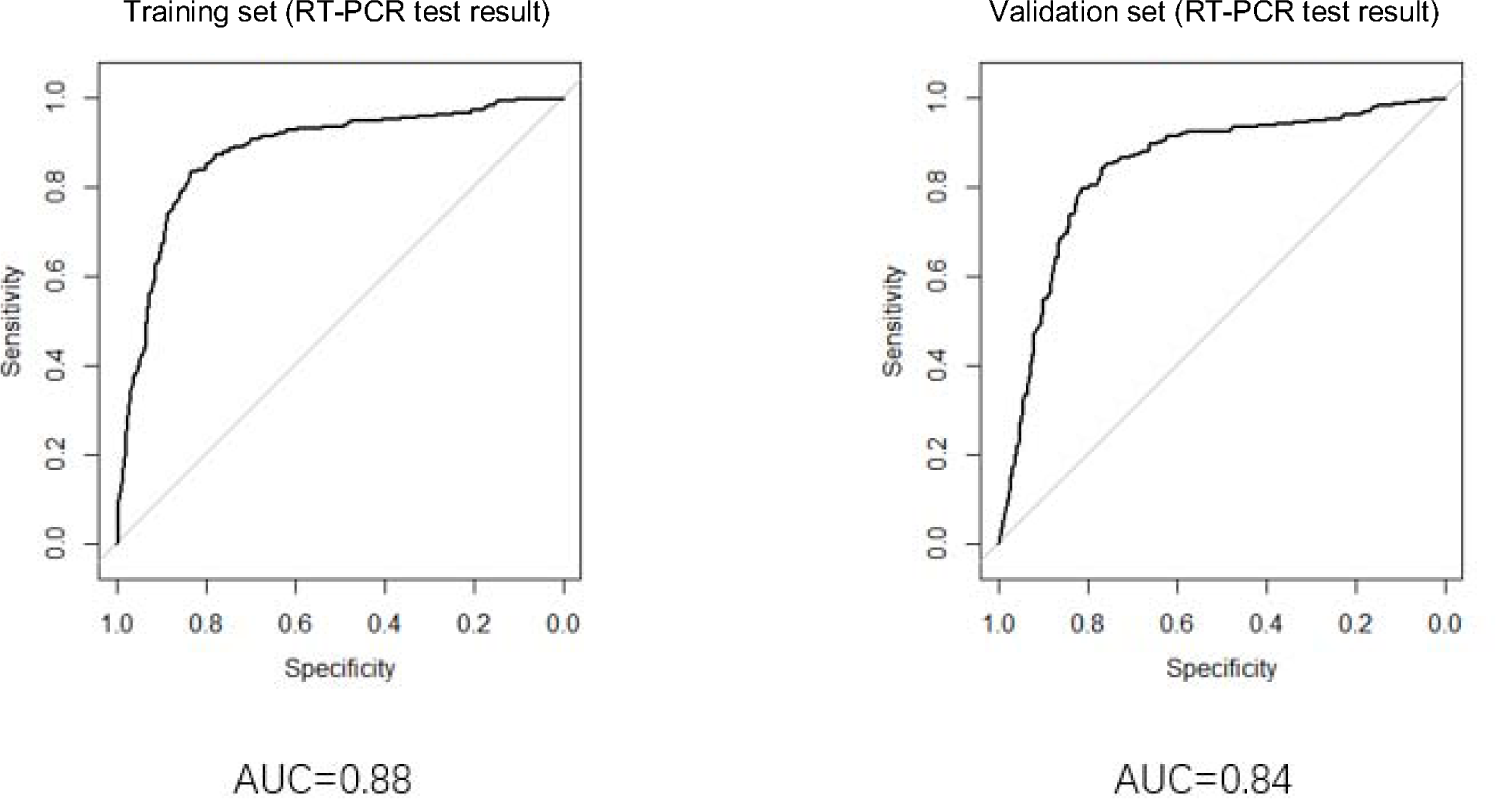
Goodness of fit of the probability model for COVID-19 in the training dataset and the validation dataset

### Comparison between the probability model and the clinical diagnosis

Figure 5 illustrates that the sensitivity of clinical diagnosis was 12.4% (17/137, 95% CI: 6.9%-17.9%) in the subset of 137 indeterminate cases who initially did not have RT-PCR tests and subsequently had positive RT-PCR results, and 17.7% (11/62, 95% CI: 8.2%-27.3%) in the subset of 62 suspected cases who initially had false-negative RT-PCR test results and subsequently had positive RT-PCR results. In comparison, the sensitivity of the probability model for COVID-19 in these two subsets were 96.4% (132/137, 95% CI: 91.7%-98.8%) and 95.2% (59/62, 95% CI: 86.5%-99.0%), respectively. The probability model demonstrated better performance in sensitivity than the clinical diagnosis, which is of statistical significance (*P*<0.05).

**Figure 5:**
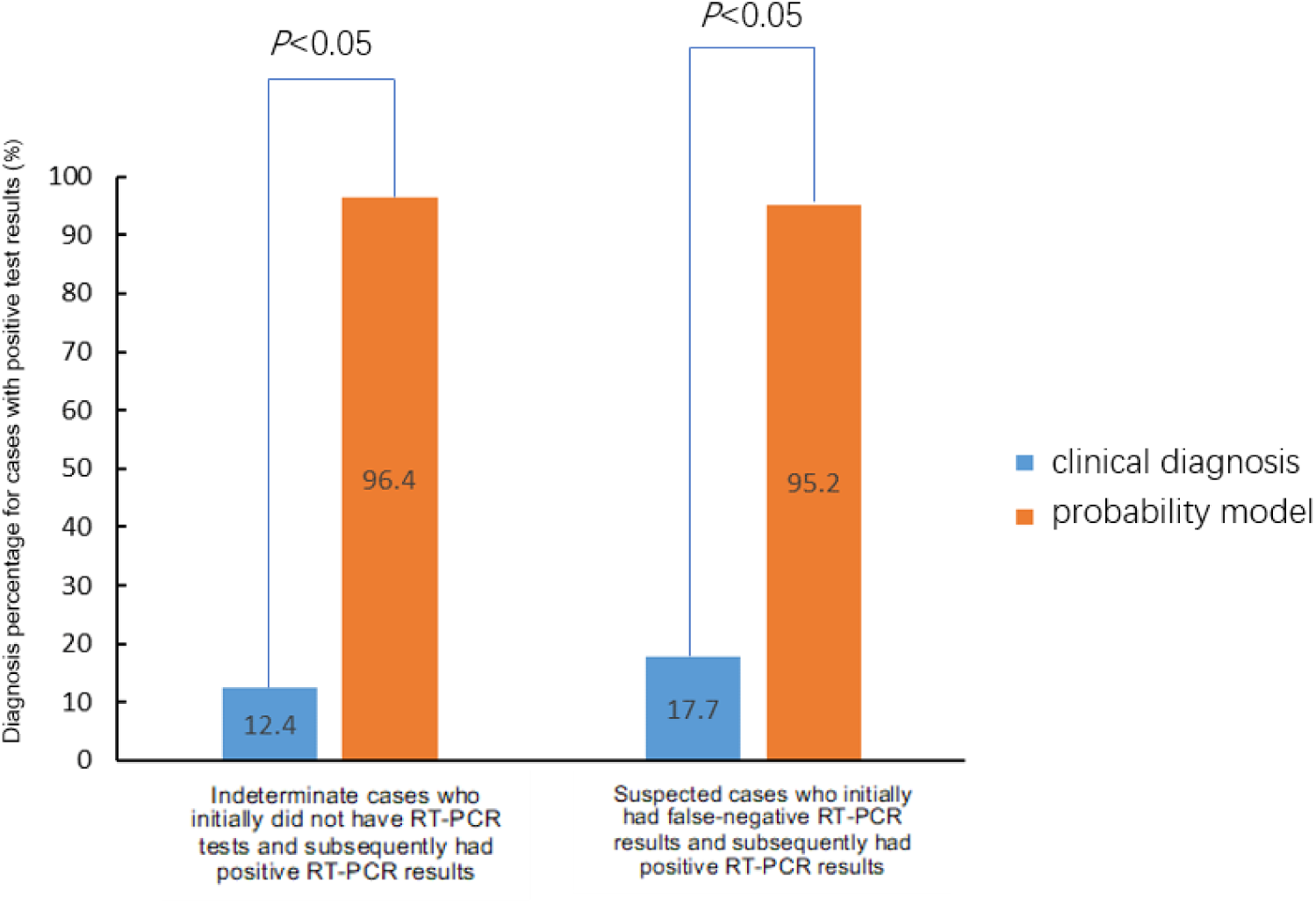
Comparison between the probability model and the clinical diagnosis

### Diagnostic value of the model in different subsets

As indicated in Figure 6, the diagnosis percentage of the model for asymptomatic patients was 83.6% (51/61, 95% CI: 74.3%-92.9%) in the validation dataset, 100% (16/16, 95% CI: 79.4%-100%) in the subset of indeterminate cases who initially did not have RT-PCR tests and subsequently had positive RT-PCR results and 71.4% (5/7, 95% CI: 29.0%-96.3%) in the subset of suspected cases who initially had false-negative RT-PCR test results and subsequently had positive RT-PCR results. Figure 7 shows that the diagnosis percentage of the model was 96.4% (132/137, 95% CI: 91.7%-98.8%) in the COVID-19 patients who initially did not have RT-PCR tests and subsequently had positive RT-PCR results; to break this down further: 96.2% (125/130, 95% CI: 91.2%-99.7%) in patients with pneumonia, and 100% (7/7, 95% CI: 76.8%-100%) in patients without pneumonia; the diagnosis percentage was 95.2% (59/62, 95% CI: 86.5%-99.0%) in the subset of COVID-19 patients who initially had false-negative RT-PCR test results and subsequently had positive RT-PCR results; to break this down further: 100% (53/53, 95% CI: 96.4%-100%) in patients with pneumonia and 66.7% (6/9, 95% CI: 29.9%-92.5%) in patients without pneumonia. The probability model is of high value in the diagnosis of different subsets of COVID-19 cases.

**Figure 6:**
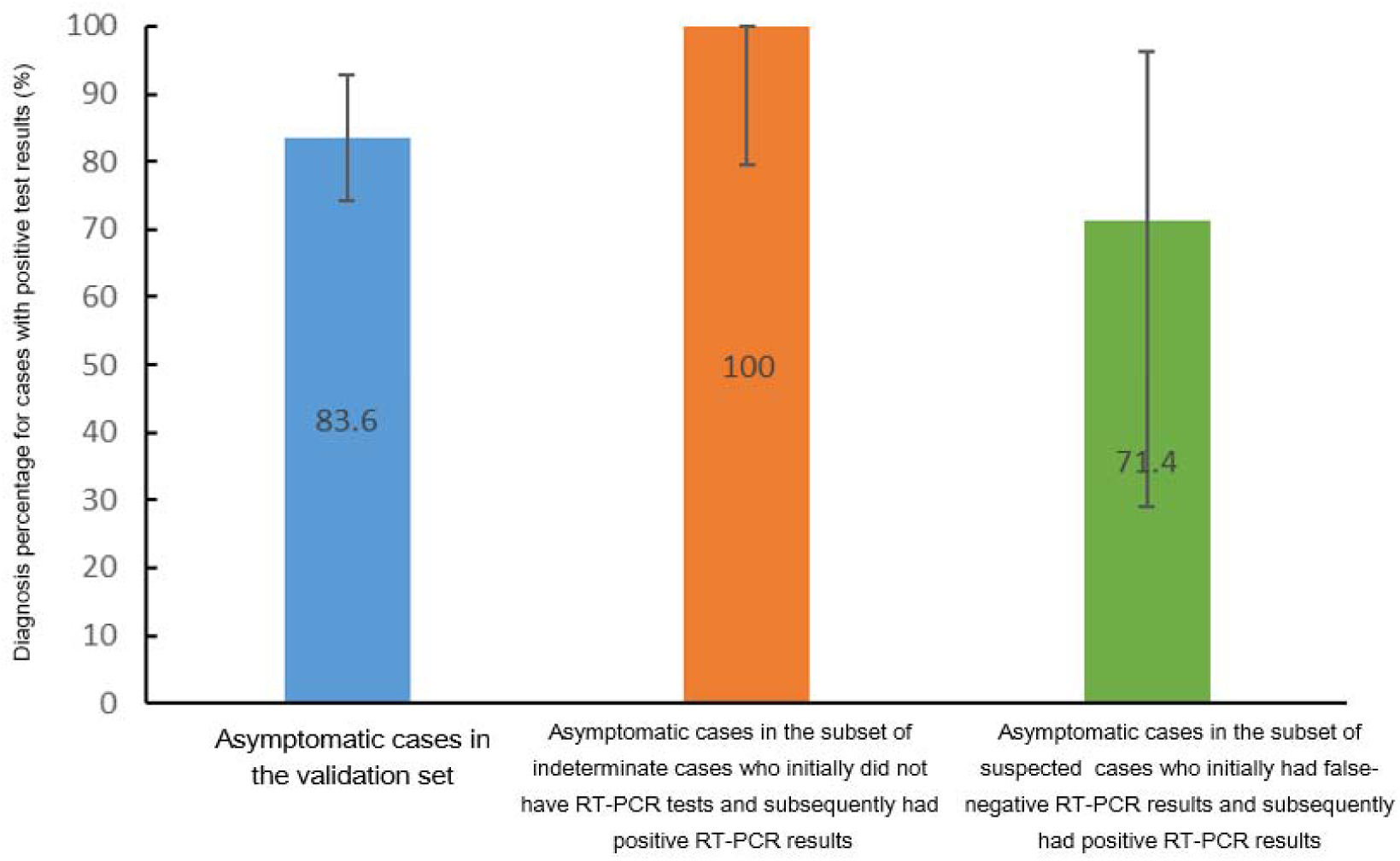
Diagnostic value of the model in asymptomatic COVID-19 patients

**Figure 7:**
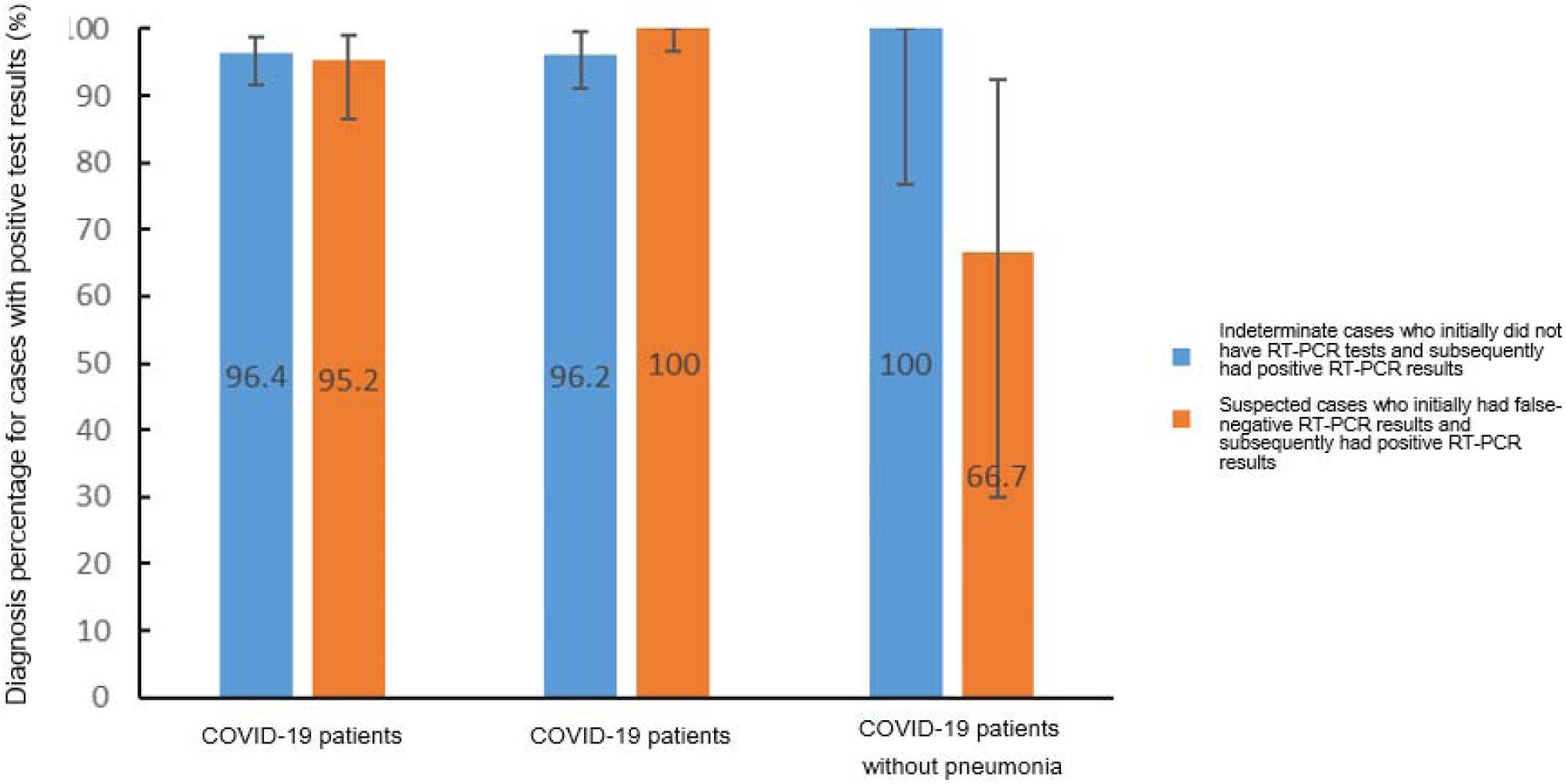
Diagnostic value of the model in COVID-19 patients with or without pneumonia

### User feedback on nCapp

We applied the ‘Questionnaire Star’ online survey for nCapp Cloud Plus Terminal to collect feedback from the terminal users, and 380 questionnaires were collected (40.6% of the total nCapp registered users). The survey covered the basic information of the user, nCapp working environment, and satisfaction. In addition, users were required to rank different platforms based on their own experience, including WeChat mini programs, mobile Apps, desktop Apps, and statistics websites or webpages. The results showed that significantly more users were ‘satisfied’ or ‘very satisfied’ with the WeChat mini program. In summary, the internet-based WeChat mini program nCapp received highly positive feedback from Chinese doctors (overall satisfaction rate, 90.9%). The ‘availability and sharing convenience of the App’ and ‘fast speed of log-in and data entry’ were the most valued features of nCapp, which had significantly better performance compared with other telemedicine tools. (Figure 8)

**Figure 8:**
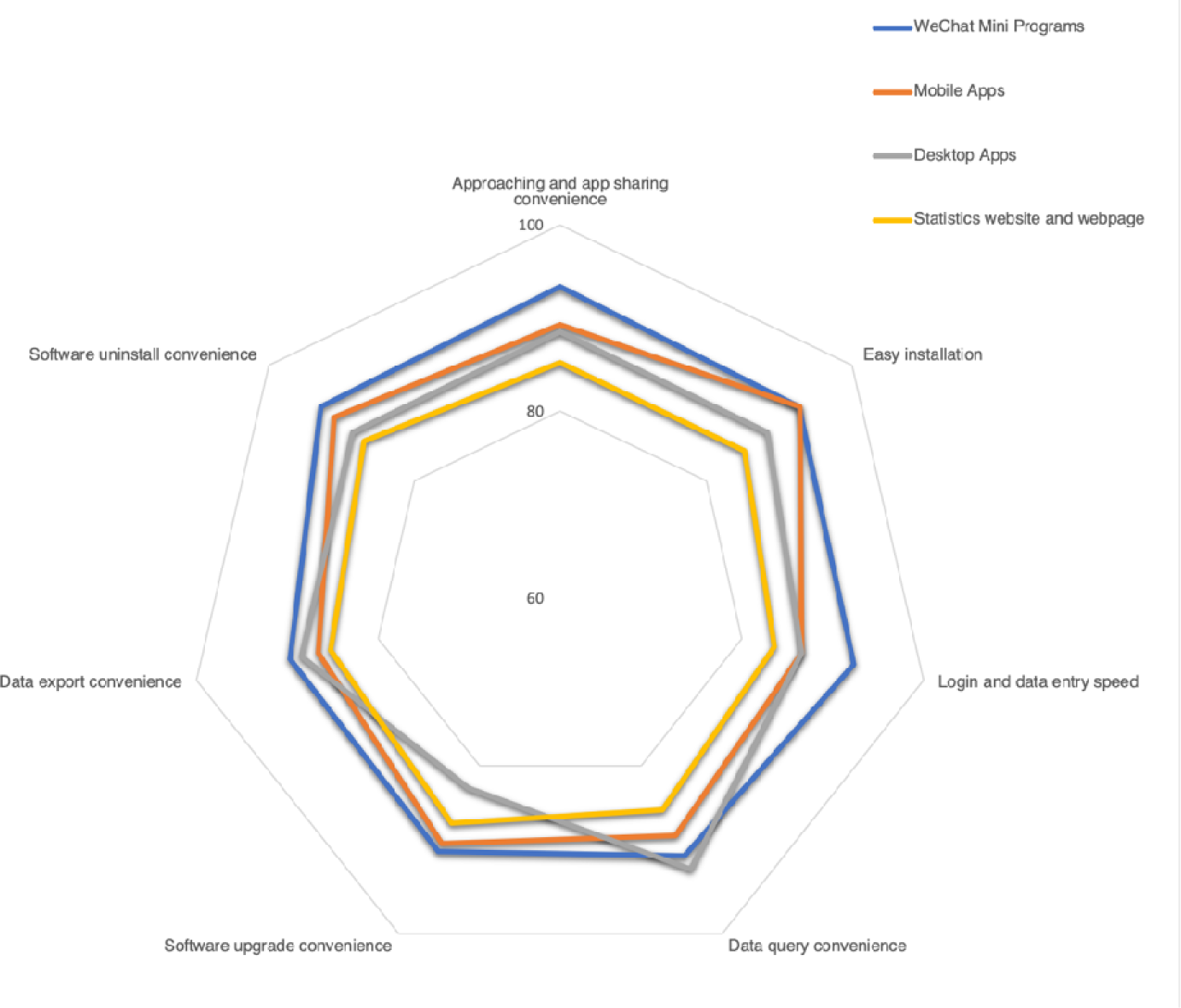
Evaluation of user experience

## Discussion

This study has demonstrated that the user-friendly mobile-based tool nCapp can quickly and easily synchronize and share clinical diagnosis and treatment information for research, that is, it helps with the optimization and screening of the determinants required for the diagnosis of COVID-19 while doctors are engaged in clinical practice.^8^ By applying classical statistical methods to optimize and select the diagnostic determinants, nCapp facilitates the identification and diagnosis of COVID-19 patients and prompts advice on better management of the cases, especially suspected cases who had initial false-negative RT-PCR test results, or indeterminate cases who initially did not have RT-PCR tests as they did not meet the criteria for suspected cases. These features of nCapp can ultimately contribute to the early detection and timely quarantine of patients. Survey results from users indicated that nCapp had significantly better performance in ‘availability and sharing convenience of the App’ and ‘fast speed of log-in and data entry’ compared with other telemedicine tools.

Currently more than 4.02 million COVID-19 cases have been reported worldwide, with a continuing trend of transmission at the community level^6^, which implies a certain number of patients with hidden infections in the epidemic areas^11^. The pathogenesis of COVID-19 is different from that of SARS or other viral diseases, and evidence shows that a patient’s viral load can last longer^12^, while cases of recurrence and secondary infection in patients who have been ‘cured’ of COVID-19 have also been reported^13^. Therefore, it is essential to identify suspected and indeterminate cases with false-negative RT-PCR results, to avoid missed diagnoses or misdiagnoses due to false-negative results, and to provide early quarantine of the infectious sources to halt the spread of the disease. It is also noteworthy that COVID-19 patients with initial false-negative RT-PCR test results are a high risk for the general population^14^. Our study has demonstrated that nCapp can reduce missed diagnoses, including in the 137 indeterminate cases and 62 suspected cases in our dataset. Additionally, COVID-19 presents challenges for health systems. There is an urgent need to train doctors (including GPs) to have a good understanding of the clinical manifestations, laboratory test results, and diagnostic criteria of COVID-19, and to enhance standardized protocols for diagnosis and treatment^15^. To address these challenges, it is necessary to simplify the current complex diagnosis-related factors and develop convenient and user-friendly intelligent assistant tools, which enable all doctors to quickly learn and apply for the timely identification, diagnosis, and management of suspicious patients.

However, the current consensus guidelines include multiple clinical manifestations and laboratory tests for the diagnosis of COVID-19. To avoid missed diagnoses and to facilitate accessibility in areas with poor medical conditions, a total of 20 factors are regarded as relevant diagnostic factors^4^. The large number of diagnostic factors becomes a barrier for doctors from different specialties to make comprehensive judgments. For less experienced doctors, it is difficult to differentiate disease-related factors and important diagnostic determinants. Therefore, it is necessary to optimize and select the key determinants that are indispensable to the diagnosis for doctors from different specialties (including GPs) in the clinical practice, thus reducing missed diagnoses. We applied the statistical method of a multi-factor linear regression model and found that the multiple diagnostic factors can be optimally reduced to 9 items, which will be more convenient for doctors to apply in clinical practice, especially with the support of nCapp. This raised another critical question of whether new problems with missed diagnoses and misdiagnoses will emerge after the optimal reduction of diagnosis-related factors to 9 items. We addressed this concern by carefully screening the model, and results showed that the diagnosis percentage of the model for asymptomatic patients was 83.6% (95% CI: 74.3%, 92.9%) in the validation set, 100% (95% CI: 79.4%, 100%) in the subset of indeterminate cases who initially did not have RT-PCR tests and subsequently had positive RT-PCR results, and 71.4% (95% CI: 29.0%, 96.3%) in the subset of suspected cases who initially had false-negative RT-PCR test results and subsequently had positive RT-PCR results. As indicated in Figure 7, the diagnosis percentage of the model was 96.4% (95% CI: 91.7%, 98.8%) in the subset of COVID-19 patients who initially did not have RT-PCR tests and subsequently had positive RT-PCR results; to break this down further: 96.2% (95% CI: 91.2%, 99.7%) in patients with pneumonia, and 100% (95% CI: 76.8%, 100%) in patients without pneumonia; the diagnosis percentage was 95.2% (95% CI: 86.5%, 99.0%) in the subset of COVID-19 patients who initially had false-negative RT-PCR test results and subsequently had positive RT-PCR results; to break this down further: 100% (95% CI: 96.4%, 100%) in patients with pneumonia, and 66.7% (95% CI: 29.9%, 92.5%) in patients without pneumonia.

Characterized by universality and versatility, nCapp can quickly and easily synchronize and share clinical diagnosis and treatment information for research, providing an accessible and convenient platform for real-world research^16^. The nCapp WeChat mini program is a new open-source cloud platform specially designed for clinical researchers. A mini program can be developed in a short time and easily accessed and distributed among WeChat users^17^. Different from traditional applications, a WeChat mini program does not need any configuration, which is of great convenience. Clinical researchers and doctors can log in to the program within a few seconds, complete the registration and electronic consent forms at the same time, and immediately start to collect first-hand information and conduct online big-data analyses by applying statistical models. This system facilitates standardized healthcare management, and users can easily implement the clinical guidelines in real time^18^ following the standardized diagnosis and treatment required by the Chinese Guideline. nCapp has advantages in its functions compared with other COVID-19-related Apps that solely collect patients’ activity information and geographic location through the smartphone’s GPS signal^19^. Survey results from ‘Questionnaire Star’ representing the feedback of users showed that nCapp integrated a standardized case reporting system with an intelligent assistant diagnosis and treatment system, which enabled it to have significantly better performance in ‘availability and sharing convenience of the App’ and ‘fast speed of log-in and data entry’ than other telemedicine tools, and to contribute to addressing the issues in the current clinical diagnosis of COVID-19. In particular, in some low- and middle-income countries, it is possible to make up for the lack of diagnostic reagents at an early stage of the outbreak by using nCapp, which features convenience and accessibility, and to reduce the risk of widespread people-to-people transmission due to the inability to identify suspicious patients timely, which is of great significance for the global control of the COVID-19 pandemic.^16^ nCapp terminal users include frontline doctors from different specialties, the staff of the Chinese Centers of Disease Control, and medical professionals at Public Health Centers in China. Through the application of the program, more confirmed, suspected, and indeterminate cases can be tested, quarantined, and managed timely to provide early alert and isolation of diagnosed or suspicious sources of infection.

One limitation of this study is that the uploaded cases were mainly moderate-to-severe COVID-19 cases, which leads to restrictions on the application of nCapp for mild patients. In addition, WeChat is not widely used in European or American countries, despite its popularity as a social media cloud platform in China and its emerging application for medical purposes. To address this issue, one approach would be to popularize WeChat, or to develop tools on a similar platform such as WhatsApp for the diagnosis and treatment of COVID-19, in compliance with HIPAA regulations to protect patient data privacy.

## Data Availability

N/A

---

**Figure S1.**
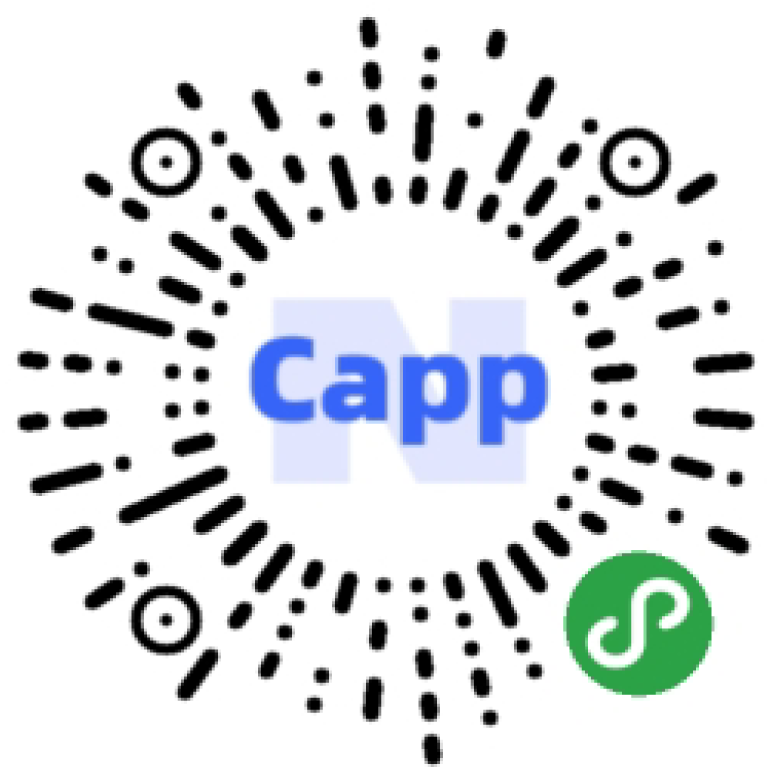
QR code for nCapp

## Notes

### Competing Interest Statement

The authors have declared no competing interest.

### Clinical Trial

NCT04275947

### Funding Statement

The study is supported by Shanghai Municipal Key Clinical Specialty(shslczdzk02201) and Shanghai Top-Priority Clinical Key Disciplines Construction Project (2017ZZ02013).

### Author Declarations

The study protocol was approved by the ethics committee of Zhongshan Hospital Fudan University (approval number B2020-032R).

